# Penetrance of *HFE* haemochromatosis variants to clinical disease: polygenic risk score associations in UK Biobank

**DOI:** 10.1101/2022.03.08.22272084

**Authors:** Luke C. Pilling, Janice L. Atkins, David Melzer

## Abstract

**Background:** The iron overload condition Hereditary Heamochromatosis (HH) can cause liver cirrhosis and cancer, diabetes and arthritis. In Europeans, most HH disease occurs in male *HFE* p.C282Y homozygotes, yet only a minority of homozygotes in the general population develop these conditions. We aimed to determine whether common genetic variants influencing iron levels or risks for liver, diabetes or arthritis diagnoses in the general population also modify clinical penetrance in *HFE* p.C282Y and p.H63D carriers.

**Methods:** 1,294 male and 1,596 female UK Biobank European-ancestry *HFE* p.C282Y homozygous participants with electronic medical records up to 14 years after baseline assessment were studied. Polygenic risk scores (PRS) quantified genetic effects on blood iron biomarkers and relevant diseases (identified in the general population). Analyses were repeated in 10,699 p.C282Y/p.H63D compound heterozygotes.

**Results:** In male p.C282Y homozygotes, higher iron PRS increased risk of liver fibrosis or cirrhosis diagnoses (top 20% of iron PRS had Odds Ratio 4.90: 95% Confidence Intervals 1.63 to 14.73, p=0.005 versus bottom 20%), liver cancer, and osteoarthritis, but not diabetes. The liver cirrhosis PRS also associated with increased liver cancer diagnoses, and greater type-2 diabetes PRS increased risk of type-2 diabetes. In female p.C282Y homozygotes, osteoarthritis PRS was associated with increased osteoarthritis diagnoses, and type-2 diabetes PRS with type-2 diabetes. However, the iron PRS was not robustly associated with diagnoses in p.C282Y homozygote females, or in other p.C282Y/p.H63D genotypes.

**Conclusions:** *HFE* p.C282Y homozygote penetrance to clinical disease in a large community cohort was partly explained by common genetic variants that influence iron and risks of related diagnoses in the general population. Including PRS in HH screening and diagnosis may help in estimating prognosis and treatment planning.

**Lay Summary:** *Two or three sentences summarizing the main message of the article expressed in plain English to describe your findings to a non-medical audience*.

- Hereditary Haemochromatosis, an iron overload condition, is the most common genetic disease in Northern Europeans; 1 in 150 people carry two copies of the highest risk mutation (called *HFE* p.C282Y).
- Only a minority of those with high risk HH variants actually develop iron overload diseases, such as liver cancer, cirrhosis, diabetes and arthritis. We tested whether known genetic variants with smaller effects on iron levels in the whole population modify risk of iron overload related disease in those with the HH high risk variants. We did similar tests for variants linked to each of the diseases caused by iron overload.
- Increased genetic risk for higher iron significantly raised the likelihood of liver fibrosis, cirrhosis and cancer in mutation carriers. In the future this information could help identify at-risk haemochromatosis patients early.

## Introduction

Hereditary haemochromatosis (HH) is a genetic condition associated with iron-overload, which in European ancestry groups is predominantly caused by the *HFE* p.C282Y homozygote mutation (>95% of cases), with some additional diagnoses in p.H63D homozygotes [1]. The p.C282Y mutation leads to reduced plasma hepcidin levels, raised ferritin and transferrin saturation levels, and a gradual accumulation of systemic iron in adults [2]. Clinical presentations of the condition include fatigue, arthropathy, diabetes, liver disease and hormone dysregulation, and the disease can progress to liver cirrhosis, liver cancer and cardiomyopathy [3]. However, penetrance to clinical symptoms or disease is limited: in the United Kingdom Biobank (UK Biobank) study - the largest community study thus far if *HFE* p.C282Y homozygotes (n=2,890) - we estimated that only 25.3% of p.C282Y homozygous males and 12.5% of homozygous females were diagnosed with haemochromatosis by age 65 [4]. These estimates were similar to a 2015 study across 7 American medical systems (eMERGE [5]: n=106 homozygotes) which reported that 24.4% of male and 14.0% of female p.C282Y homozygotes were diagnosed with haemochromatosis (mean age 66.4 years ±15.8), with Kaplan Myer survival curves suggesting 50% of the homozygote men and 25% of homozygote women eventually diagnosed with haemochromatosis by the age of 90.

This limited clinical penetrance may be explained in part by environmental factors [6], including high alcohol consumption and hepatitis C virus infection for liver fibrosis or cirrhosis, but there is also evidence for genetic factors being involved (Radio et al, 2015). For example, in a genome-wide association studies (GWAS) in 474 unrelated p.C282Y homozygotes, single nucleotide polymorphism (SNP) rs3811647 in the *TF* (Transferrin) gene was associated with serum iron but not clinical phenotypes [7]. It explained 7.7% of the variance of serum transferrin concentration and 4.7% of the variance of serum iron levels. We previously reported preliminary evidence that common variants affecting iron levels may interact with p.C282Y genotype to increase risk of disease [8], but more evidence is required to understand the modifying effects of common variants on p.C282Y penetrance.

HH appears to meet several criteria for genetic screening [9], but the low clinical penetrance in community p.C282Y homozygotes was a major factor in limiting screening to close relatives. A better understanding of the limited penetrance might improve prediction of prognosis and might allow more targeted screening for those at risk of disease. We therefore aimed to identify whether common variants linked to variation in iron measures are associated with a clinical diagnosis of HH and related outcomes (especially liver fibrosis or cirrhosis) within *HFE* p.C282Y homozygotes, and separately in other *HFE* genotype groups. We used the UK Biobank, a cohort of over 500,000 community volunteers receiving routine clinical care: the UK Biobank consent procedure explicitly involved no personal feedback of genetic findings.

## Methods

The UK Biobank study (UKB) includes data on 502,634 volunteers aged 40 to 70 at baseline study invitation. Recruitment was through postal invitation to people registered with the UK National Health Service, living within 25 miles of 22 assessment centres in England, Scotland and Wales. Participants consented to genotyping, and for data linkage for follow-up by hospital admission medical records (hospital episode statistics, HES), cancer registry, primary care (general practice, GP), and death certificates. UKB volunteers tended to be healthier at baseline than the general UK population [10].

Data are available to any bone fide researcher following application to UKB (www.ukbiobank.ac.uk/register-apply). The North West Multi-Centre Research Ethics Committee approved the collection and use of UK Biobank data (Research Ethics Committee reference 11/NW/0382). Access to UKB was granted under Application Number 14631.

### Disease ascertainment

Disease ascertainment was by subject responses to questionnaire items on doctor diagnosed diseases at baseline (2006-2010), combined with International Classification of Diseases 10^th^ revision (ICD-10) coded hospital inpatient records, cancer registry data, and read codes from primary care data which was available for approximately 45% of the participants. The censoring dates for the three sources of electronic medical records were up to September 2021 for HES (England; July 2021 for Scotland; Feb 2018 for Wales), July 2019 for cancer register (England and Wales; October 2015 for Scotland), and August 2017 for primary care (Wales; March 2017 for Scotland; May 2017 for England Vision supplier and August 2016 for England TPP supplier). Diagnoses ascertained were haemochromatosis (ICD-10 code E83.1), liver conditions (any, based on ICD-10 codes K70-77), liver fibrosis or cirrhosis (ICD-10 codes K74*), liver cancer (ICD-10 C22*), diabetes mellitus (predominantly diagnosed as type 2 (E11*) but including type 1 (E10*), as typing of diabetes in haemochromatosis may be unclear), and osteoarthritis (ICD-10 codes M15.0; M15.1; M15.2; M15.9; M16.0; M16.1; M17.0; M17.1; M18.0; M18.1; M19.0). Corresponding primary care diagnosis codes were identified using the UK Biobank “Clinical coding classification systems and maps” resource to map ICD-10 codes to Read2/CTV3 (https://biobank.ctsu.ox.ac.uk/crystal/refer.cgi?id=592).

### Genotyping in UK Biobank

Participants were genotyped using two almost identical (>95% shared variants, n=805,426 total) microarray platforms: the Affymetrix Axiom UK Biobank array (in 438,427 participants) and the Affymetrix UKBiLEVE array (in 49,950 participants). The central UKB team performed genotype imputation in 487,442 participants, increasing the number of genetic variants to ∼96 million [11]. Because *HFE* p.C282Y is largely restricted to Europeans, we analysed 451,427 (93%) participants who self-reported as “white European” and were confirmed as of genetically European ancestry (described [12]). 445,521 participants (98.7% of 451,427) had *HFE* p.C282Y (rs1800562) imputed with 100% confidence and 5,723 were recoded (i.e. estimated genotype dose between 0 and 0.25 set to 0, values between 0.75 and 1.25 set to 1, and finally between 1.75 and 2 to 2); 183 participants (0.04%) were excluded due to imprecise imputation, yielding 451,243 participants in analyses. *HFE* p.H63D (rs1799945) was directly genotyped on the microarray.

### Polygenic risk scores for iron status biomarkers

We created Polygenic Risk Scores (PRS) for four iron status biomarkers using 128 non-*HFE* variants (p.C282Y/p.H63D variants excluded) identified in a GWAS of 257,953 individuals [13]. We used 20 variants associated with iron itself, 64 associated with ferritin, 19 with transferrin saturation, and 41 with total iron-binding capacity (Supplementary Table 1). We excluded a small number of variants identified in the original GWAS if they were not present in the UK Biobank imputed data (v3), if the minor allele frequency was <0.1%, if there was significant deviation from Hardy-Weinberg equilibrium (p<5*10-8), or if the imputation quality (INFO score) was below 80% (see Supplementary Table 2 for details). For each participant, the number of trait-raising alleles was counted, weighted by the effect size reported in the published GWAS [13] (effects and effect alleles reported in Supplementary Table 1; excluded SNPs with criteria are listed in Supplementary Table 2).

### Polygenic risk scores for heamochromatosis-associated co-morbidities

Genetic variants associated with liver cirrhosis, osteoarthritis, type-1 diabetes (T1D) and type-2 diabetes (T2D) in general population studies (i.e. not specific to haemochromatosis) were identified from published GWAS [14]–[17], and for each participant the number of trait-raising alleles was counted, weighted by the effect size reported in the published GWAS. A small number of variants identified in the original GWAS were excluded if: the SNP was +/- 250kb of p.C282Y, they were not present in the UK Biobank imputed data (v3), if the minor allele frequency was <0.1%, if there was significant deviation from Hardy-Weinberg equilibrium (p<5*10^−8^), or if the imputation quality (INFO score) was below 80%.

### Missing data

We excluded participants without imputed genotype data (n=15,233/502,642, 3.03%), those with imprecise imputation for p.C282Y (n=183/487,409, 0.037%), and those who had withdrawn from the study at the time of analysis (December 2021). Less than 0.5% of participants had no answers to questions on self-reported diseases. Given the low level of missing data, we excluded participants with missing data from individual analysis, as needed.

### Statistical analysis

PRS represent life-long predisposition to higher/lower levels of iron status biomarkers; we therefore used logistic regression models to test the hypothesis that “PRS for trait X is associated with ever bring diagnosed with outcome Y,” adjusted for age at end of medical records follow up, assessment center, and genetic principal components of ancestry 1 to 10.

To adjust for multiple statistical testing and reduce the false discovery rate we used the Benjamini-Hochberg method to identify p-values <0.05 after multiple testing correction. These are indicated on Figures and in the text.

We applied two-sample Mendelian randomization (MR) methods to test the robustness of the associations seen using the two-stage least squares approach (i.e. the one-sample approach testing associations between iron PRS outcomes within UKB). We used the R package ‘RadialMR’ to test for significant pleiotropy (using the MR Egger approach [18]) and significant outliers or heterogeneity in the variant effects (Radial MR approach) [19].

## Results

In 2,890 *HFE* p.C282Y homozygotes in UK Biobank European-ancestry participants we identified 771 (26.7%) with a haemochromatosis diagnosis at the end of available electronic medical record data (HES up to Sept 2021, or GP data up to Sept 2017; GP data available in 45% of cohort). Diagnosis was more common in males (33.2% of 1,294 males vs. 21.4% of 1,596 females) and mean age at diagnosis was 61.5 years (60.1 in males). See Table 1. In non-p.C282Y homozygotes diagnosis was substantially less common (703 diagnoses in 448,441 participants), as expected (see Supplementary Table 3 for details including number of comorbidity diagnoses in other *HFE* genotype groups).

**Table 1:** Characteristics of the UK Biobank *HFE* p.C282Y homozygote participants of European ancestry, stratified by sex.

|  | All |  | Males |  | Females |  |
| --- | --- | --- | --- | --- | --- | --- |
|  | N | % | N | % (M) | N | % (F) |
| *HFE* p.C282Y homozygotes | 2,890 |  | 1,294 |  | 1,596 |  |
| Diagnosis*: Haemochromatosis | 771 | 26.7 | 430 | 33.2 | 341 | 21.4 |
| Liver disease (any) | 220 | 7.6 | 141 | 10.9 | 79 | 4.9 |
| - Liver fibrosis or cirrhosis | 78 | 2.7 | 60 | 4.6 | 18 | 1.1 |
| - Liver cancer | 30 | 1.0 | 27 | 2.1 | 3 | 0.2 |
| Osteoarthritis | 721 | 24.9 | 326 | 25.2 | 395 | 24.7 |
| Type-1 Diabetes | 45 | 1.6 | 32 | 2.5 | 13 | 0.8 |
| Type-2 Diabetes | 266 | 9.2 | 163 | 12.6 | 103 | 6.5 |
| Any HH co-morbidity^ | 1,017 | 35.2 | 507 | 39.2 | 510 | 32.0 |
|  | **Mean** | **SD** | **Mean** | **SD** | **Mean** | **SD** |
| Age (end follow-up or death) | 69.6 | 8.0 | 69.3 | 8.1 | 69.8 | 8.0 |
| Age at haemochromatosis diagnosis | 61.5 | 9.3 | 60.1 | 9.7 | 63.2 | 8.5 |

\* Diagnosis ever recorded in data from baseline self-report, HES up to Sept 2021, Cancer registry up to July 2019, or GP data up to Sept 2017 (GP data available in 45% of cohort).
^ any diagnosis of liver disease, osteoarthritis, type-1 diabetes or type-2 diabetes. HH = hereditary haemochromatosis. SD=standard deviation.

### Polygenic risk score of common iron-increasing genetic variants affects penetrance in *HFE* p.C282Y homozygous participants

We tested associations between polygenic risk scores (PRS) for four blood iron status biomarkers [13] and haemochromatosis-associated comorbidities in male *HFE* p.C282Y homozygotes (Figure 1).

**Figure 1:**
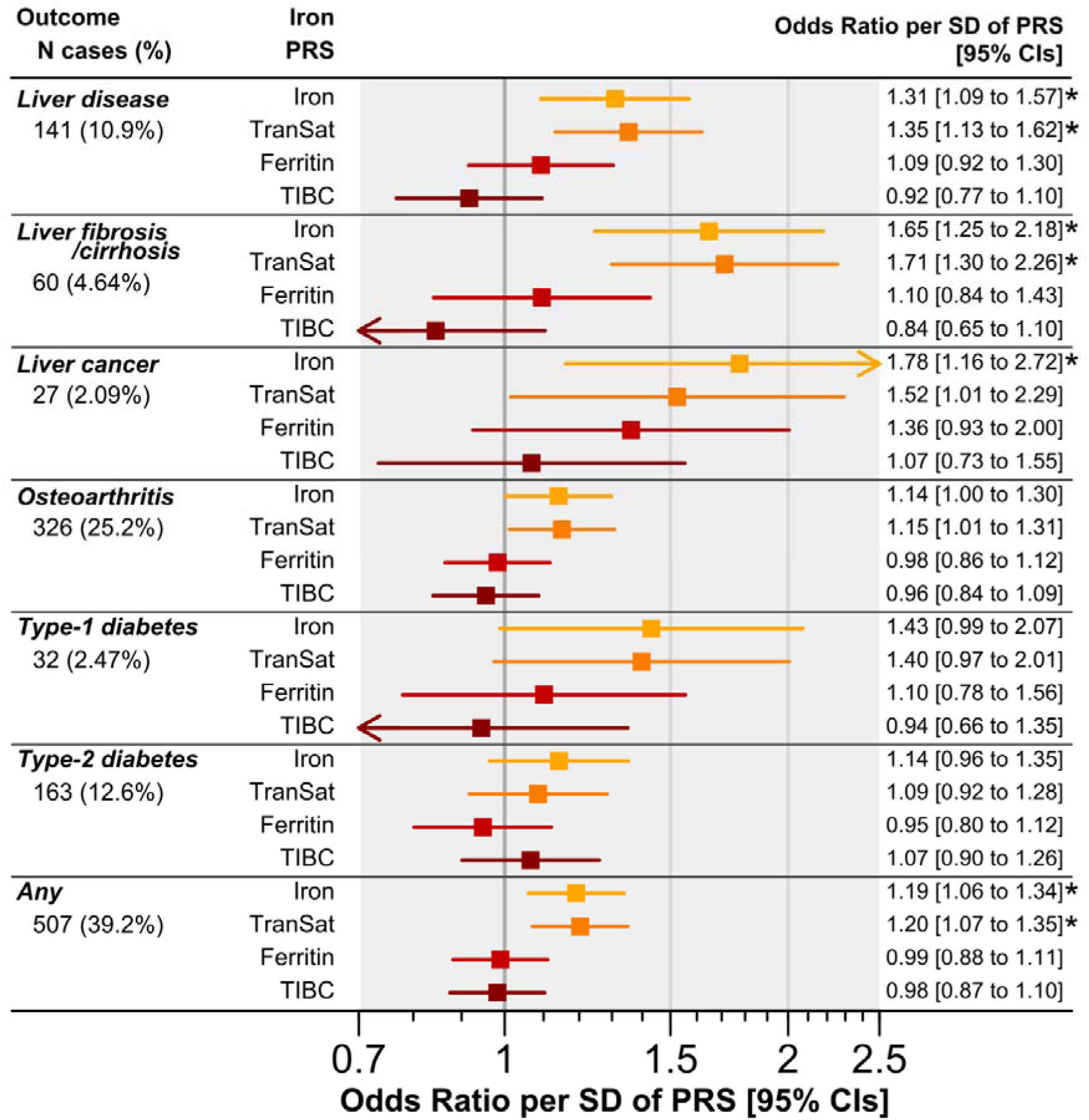
Linear associations between four iron status biomarker PRS and HH co-morbidities in *HFE* p.C282Y homozygous males. Results are from logistic regression models adjusted for age, assessment center, and principal components of ancestry 1 to 10. % = percentage of 1,294 *HFE* p.C282Y homozygous males of European genetic ancestry who ever received a diagnosis in the available data (up to Sept. 2021). HH = hereditary haemochromatosis. Iron PRS = polygenic risk score for the iron status biomarker (TranSat = transferrin saturation, TIBC = total iron-binding capacity). CI = confidence interval. SD = standard deviation. Outcomes: Liver disease includes any related diagnosis. Liver cirrhosis is specifically cirrhosis or fibrosis diagnoses. “Any” = any diagnosis of liver disease, osteoarthritis, type-1 diabetes or type-2 diabetes. See Supplementary Table 4 for details. * = false discovery rate adjusted p-value <0.05 (Benjamini-Hochberg method).

Iron PRS was associated with increases in likelihood of ever being diagnosed with related diseases, especially liver fibrosis or cirrhosis (Odds Ratio per Standard Deviation increase in Iron PRS 1.65 95% Confidence Intervals 1.25 to 2.18, p=5*10^−4^), liver cancer (OR 1.69: 95%CI 1.01 to 2.81, p=0.04), or any liver disease (OR 1.31: 95%CI 1.09 to 1.57, p=0.003) in logistic regression models adjusted for age, assessment center, and principal components of ancestry 1-10 (Supplementary Table 4), after adjustment for multiple testing. Iron PRS was also nominally associated with increased likelihood of osteoarthritis (OR 1.14: 95%CI 1.00 to 1.30, p=0.046), but trends with T1D (OR 1.43: 95%CI 0.99 to 2.07, p=0.058) or T2D (OR 1.14: 95%CI 0.96 to 1.35, p=0.12) did not reach significance. Iron PRS was also significantly associated with greater likelihood of ever receiving a haemochromatosis diagnosis (OR 1.33: 95%CI 1.18 to 1.50, p=3*10^−6^). In female *HFE* p.C282Y homozygotes, iron PRS increased likelihood of haemochromatosis diagnosis (OR 1.32: 95%CI 1.17 to 1.49, p=1*10^−5^) but was not associated with any comorbidities tested (p>0.05; Supplementary Table 4).

We also created a PRS for transferrin saturation using 19 genetic variants, and results were highly similar to the iron PRS results reported above (Figure 1; Supplementary Table 4), however PRS for ferritin and total iron binding capacity were not associated with diagnosis of haemochromatosis or any comorbidities (p>0.05; Figure 1; Supplementary Table 4).

To explore the association between iron PRS and diagnosis of liver fibrosis or cirrhosis further, we stratified the 1,294 male *HFE* p.C282Y homozygotes into 5 equally sized groups (quintiles) based on their iron PRS. Those into the top 20% of iron PRS (n=259) had substantially higher likelihood of being diagnosed with liver fibrosis or cirrhosis (n=19) compared to those in the bottom 20% of iron PRS (n=4 diagnoses in 259 participants) (OR 4.90: 95%CI 1.63 to 14.73, p=0.005) (Figure 2; Supplementary Table 5).

**Figure 2:**
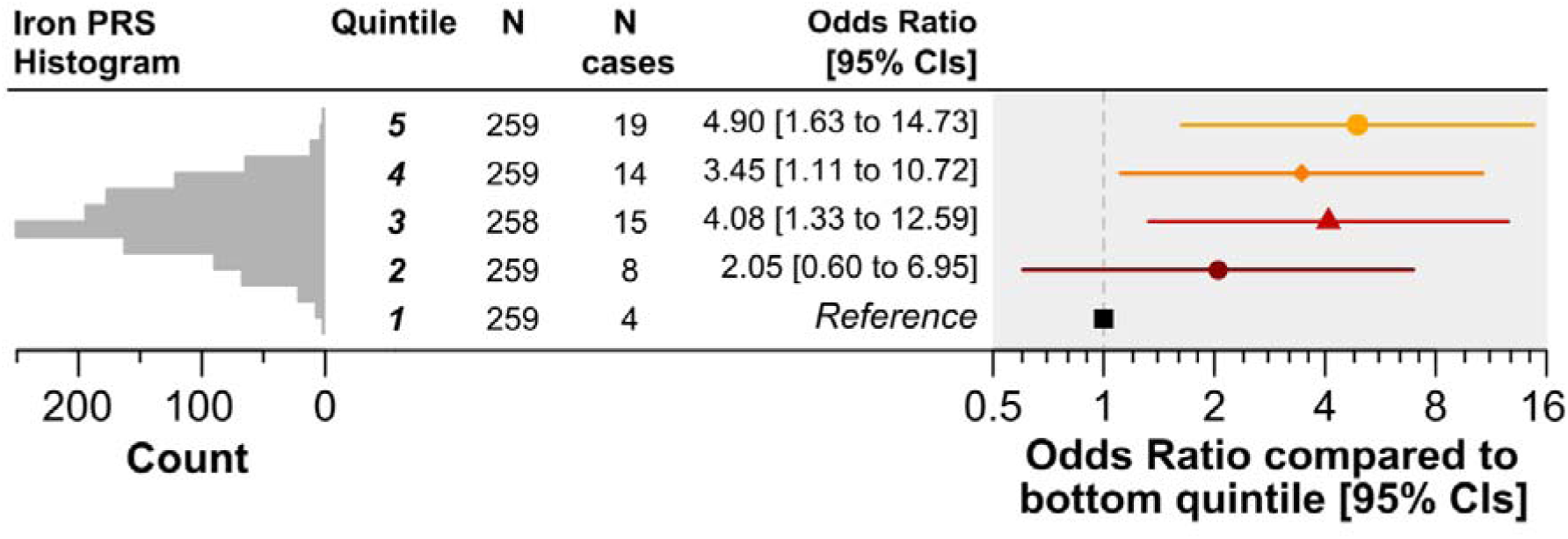
Iron PRS quintile association with diagnosis of liver fibrosis or cirrhosis in *HFE* p.C282Y homozygous males. Iron PRS is stratified into 5 equally sized groups (quintiles). Results are from logistic regression models adjusted for age, assessment center, and principal components of ancestry 1 to 10. N = number of *HFE* p.C282Y homozygous males of European genetic ancestry in quintile. N cases = number in quintile who ever received a diagnosis of liver fibrosis or cirrhosis in the available data (up to Sept. 2021). Iron PRS = polygenic risk score for total iron levels. CIs = Confidence Intervals. See Supplementary Table 5 for details including PRS cut points.

### Co-morbidity polygenic risk score associations in *HFE* p.C282Y homozygotes

Within *HFE* p.C282Y homozygous males, a PRS for liver cirrhosis was nominally associated with increased risk of liver cancer (OR 1.48: 95%CI 1.03 to 2.12, p=0.04) (Figure 3; Supplementary Table 6). The association was not significantly different to that in the *HFE* wild-type group (no p.C282Y or p.H63D genotypes) when an interaction term was included between cirrhosis PRS and *HFE* genotype (p>0.05). A PRS for osteoarthritis was significantly associated with diagnosis of osteoarthritis in *HFE* p.C282Y homozygous females (OR 1.29: 95%CI 1.14 to 1.45, p=4*10^−5^) but not males (OR 1.12: 95%CI 0.98 to 1.27, p=0.1). The association in p.C282Y homozygous females was significantly greater than that in *HFE* wild-type participants (interaction p=0.012). A PRS for T2D was significantly associated with increased likelihood of T2D diagnosis in both p.C282Y homozygous males (OR 1.86: 95%CI 1.55 to 2.24, p=2*10^−11^) and females (OR 1.72: 95%CI 1.39 to 2.12, p=6*10^−7^), though in both cases the association did not significantly differ from that seen in *HFE* wild-type genotype participants (interaction p>0.05). PRS for liver cirrhosis was not associated with diagnosis of liver cirrhosis in p.C282Y homozygotes (p>0.05) though the association was significant in the larger *HFE* wild-type group (OR_males_ 1.20: 95%CI 1.16 to 1.23, p=6*10^−38^). A T1D PRS was not significantly associated with T1D diagnosis in p.C282Y homozygotes (p>0.05), though the association was significant in *HFE* wild-type participants (OR_males_ 1.44: 95%CI 1.35 to 1.53, p=1*10^−31^).

**Figure 3:**
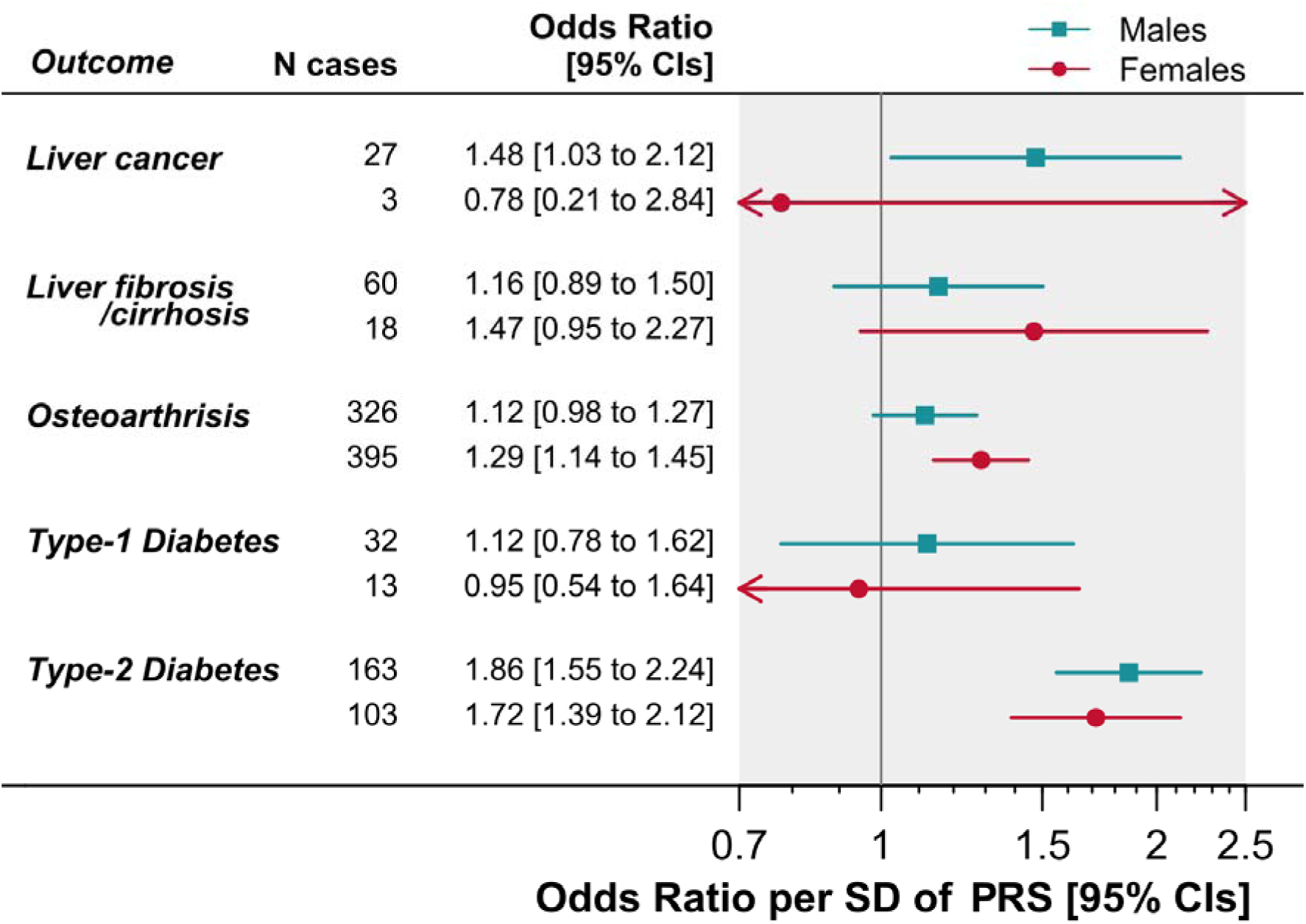
Linear increases in PRS for HH co-morbidities affect likelihood of corresponding diagnosis in *HFE* p.C282Y homozygotes. Results are from logistic regression models adjusted for age, assessment center, and principal components of ancestry 1 to 10. Liver cirrhosis PRS was tested against diagnoses of liver cancer and separately diagnoses of liver fibrosis or cirrhosis. Osteoarthritis PRS was tested against diagnoses of osteoarthritis; type-1 diabetes (T1D) PRS was tested against T1D; and type-2 diabetes against T2D. HH = hereditary haemochromatosis. PRS = polygenic risk score. CI = confidence interval. SD = standard deviation. See Supplementary Table 6 for details.

### Iron-increasing genetic variants and haemochromatosis comorbidities in other *HFE* genotype groups

Iron PRS was not associated with diagnosis of haemochromatosis comorbidities in male p.C282Y heterozygotes or p.C282Y/p.H63D compound heterozygotes (Supplementary Figure 1; Supplementary Table 4). Iron PRS was nominally associated with increased likelihood of liver fibrosis/cirrhosis in p.H63D heterozygotes (OR per SD of PRS 1.14: 95%CI 1.03 to 1.27, p=0.02), and separately with liver cancer in p.H63D homozygotes (OR 1.69: 95%CI 1.01 to 2.81, p=0.04), though neither were significant after adjustment for multiple statistical testing. Iron PRS significantly increased likelihood of diagnosis of haemochromatosis itself in p.C282Y heterozygotes (OR 1.24: 95%CI 1.08 to 1.42, p=0.002), and p.H63D homozygotes (OR 1.80: 95%CI 1.26 to 2.57, p=0.001), but not p.C282Y/p.H63D compound heterozygotes or p.H63D heterozygotes (p>0.05) (Supplementary Figure 1; Supplementary Table 4).

### Sensitivity analysis

The primary analysis in UK Biobank included all male participants of European ancestry who were homozygous for *HFE* p.C282Y (*n*=1,294): in sensitivity analysis we identified 13 pairs of participants related to the 3^rd^-degree or closer (using KING kinship analysis [20]). We randomly excluded one of each pair of related participants and repeated the primary analysis of iron PRS associations with HH comorbidities in unrelated *HFE* p.C282Y homozygous European males. The associations between iron PRS and outcomes remained consistent, suggesting the result was not biased by inclusion of related participants (Supplementary Table 7).

We repeated the primary analysis of iron PRS associations with diagnosis of liver fibrosis/cirrhosis and separately liver cancer using two-sample MR methods. We found no evidence for pleiotropy (MR Egger intercept p-values >0.05) or bias due to outliers (see Supplementary Table 8 for results), suggesting the primary analysis results presented are robust.

## Discussion

In population studies, *HFE* p.C282Y homozygosity is associated with high biochemical penetrance (to raised iron measures) but low penetrance to haemochromatosis related clinical diagnoses [6], [21]. Many genetic variants are known to influence iron measures in the general population, and risk of liver disease, arthritis or diabetes: although most individual effects are small, the cumulative expected effects of risk alleles can be computed into polygenetic risk scores (PRS) for each study participant. We therefore tested whether these PRS could explain some of the variance in clinical penetrance with the high risk *HFE* p.C282Y homozygous group. We found that carrying a greater number of common genetic variants increasing serum iron and transferrin saturation levels increased incidence of hereditary haemochromatosis (HH)-associated diseases in 2,890 *HFE* p.C282Y homozygotes and other *HFE* genotypes, in the UK Biobank, the largest community study thus far of p.C282Y homozygotes (n=2,890). We also found that p.C282Y homozygotes with high polygenic risk for liver cirrhosis, osteoarthritis, or diabetes, were more likely to develop those specific co-morbidities. Our results support the conclusion that the variable clinical penetrance of HH seen in *HFE* p.C282Y homozygotes is partly attributable to burden of polygenic risk for higher iron and higher risk of co-morbidities.

A recent GWAS meta-analysis of iron status biomarkers (irrespective of *HFE* genotype) in 257,953 individuals identified 127 loci [13]. This included loci with well-established roles in iron homeostasis and metabolism, such as *TF* (transferrin), *SLC40A1* (ferroportin-1), and *TMPRSS6* (transmembrane serine protease 6). The effect sizes for non-*HFE* variants are modest. Several previous candidate gene studies have investigated the role of modifying genetic variants amongst iron-metabolism genes in haemochromatosis (such as [22], [23]), yet sample size was a limitation. In UK Biobank we were able to extend these studies and use polygenic risk score (PRS) and Mendelian randomization (MR) methods to robustly model the cumulative risk of many small-effect genetic variants.

Higher PRS for serum iron and for transferrin saturation increased risk of liver disease, especially liver fibrosis or cirrhosis, and liver cancer. Progressive increases in serum levels are markers of increased iron absorption and are amongst the earliest signs of haemochromatosis [24]. Progressively increasing serum ferritin (hyperferritinemia) is also characteristic of haemochromatosis [24], reflecting increasing iron storage; although ferritin levels are raised in several conditions including acute inflammation, which may have resulted in the weaker association trends between higher PRS for ferritin (or total iron-binding capacity) and increase risk of liver disease or co-morbidities. Raised serum iron biomarkers are reported in other *HFE* genotype groups, especially in p.C282Y/p.H63D compound heterozygotes, compared to non-carriers [25], however evidence for the impact on clinical diagnosis and morbidity is variable [26]. We found that genetic predisposition to higher serum iron did not increase risk of haemochromatosis-associated comorbidity in these other *HFE* genotype groups.

Hepcidin is the key hormone regulating iron absorption by binding to ferroportin, limiting the release on iron into the blood [27]. *HFE* mutations result in reduced hepcidin expression in the liver and thus increased iron absorption. Included in the iron PRS are variants mapped to genes known to regulate the hepcidin cascade, such as transmembrane serine protease 6 (*TMPRSS6*), a liver-specific transmembrane protein that increases hepcidin production [28]. That no variants were identified in the hepcidin gene itself (*HAMP*) supports the hypothesis that for most patients it is the cascade events upstream of hepcidin (starting with *HFE*) that leads to hepcidin dysregulation and iron overload [29]. We saw no significant outlier variants in the MR analysis, confirming that the PRS results were not driven by a small number of effects in key genes such as *TMPRSS6*, but rather are the average effect of all iron- increasing variants.

Others have suggested a multifactorial model of hereditary haemochromatosis, characterized principally by variants in *HFE* with modifying effects of genetic and environmental factors that are yet to be fully determined [23]. Environmental factors such as alcohol consumption and hepatitis C virus infection appear to increase susceptibility to iron overload, with roles for insulin resistance, fatty acid accumulation, and ineffective erythropoiesis [30]. Protective factors are also reported, including a correct diet and positive attitude to blood donations [31]. Incomplete clinical penetrance is partly explained by these factors, and yet there is also incomplete biochemical penetrance within *HFE* genotype groups: the Hemochromatosis and Iron Overload Screening (HEIRS) Study reported that although in undiagnosed male p.C282Y homozygotes mean transferrin saturation was 76% compared to 32% in males without *HFE* mutations, there were still 16% of p.C282Y homozygotes men with transferrin saturation below 50% [25]. The PRS for transferrin saturation is reported to explain 11% of variance in transferrin saturation (although, this PRS included *HFE* variants) in 56,664 participants from the Trøndelag Health (HUNT) study, strongly supporting the hypothesis that common non-*HFE* variants modify biochemical penetrance, and our results support that this impacts penetrance to clinical disease, especially in the liver.

Limitations of this analysis include that UK Biobank volunteers tended to be healthier than the general population [10] at baseline, although this effect may have diminished during the long observed electronic medical records follow-up of over 14 years. Though hospital inpatient diagnoses were available for all participants, primary care data were only available in approximately 45% of the cohort, so estimates of penetrance to disease may be underestimated. It is possible that response rates to UK Biobank may have been affected by *HFE* mutation status or associated morbidity, but as previously reported the overall prevalence of p.C282Y homozygosity (1 in 156) was very similar to previous reports for groups of British or Irish descent [8], and the p.C282Y variant was in Hardy-Weinberg equilibrium (p>0.05) in UK Biobank, implying that the observed genotypes are present in the expected proportions, with no sign of differential loss or excess of p.C282Y homozygotes. The UK Biobank sample included a wide range of exposures and socioeconomically diverse groups [10], and prospective analyses are less affected by sample response patterns at baseline. These factors suggest that our results are robust and likely to be applicable to the UK and other European descent populations. Though iron status biomarkers were not measured in UK Biobank, biochemical penetrance of *HFE* mutations is well documented by the HEIRS study among others [25], and the effect of identified variants is reliably reported by the HUNT meta-analysis [13].

Strengths of our analysis include that UK Biobank is the largest community genotyped study of p.C282Y homozygotes (nearly 10 times bigger than HEIRS [25]). We had good ascertainment of clinical diagnoses through primary care electronic medical records and hospital admission data, though as noted above penetrance may be underestimated as primary care data is only available for ∼45% of participants. Very few p.C282Y homozygotes were diagnosed with haemochromatosis at baseline (12% of males) [8], and participants consented to not be told about UK Biobank ascertained genotypes, so results are similar to what might be expected from community screening.

The UK Biobank sample included some sets of related individuals, as assessed through genome wide variant similarity (KING kinship coefficient). In sensitivity analysis excluding one of each pair of participants related to the third-degree or closer the results were unaffected. Unfortunately, there are no data in UK Biobank on whether each related or unrelated UK Biobank participant was from a family with a strong history of haemochromatosis diagnoses, or not. Current screening focusses on families i.e., first-degree relatives of p.C282Y homozygotes, though this only identifies a minority of homozygotes; an Australian study estimated that only 2.9% of male homozygotes and 2.0% of females were identified in family screening [32]. Our results show that family relatedness didn’t affect associations, supporting calls for family-agnostic screening approaches.

## Conclusion

Overall, our findings show that *HFE* p.C282Y homozygote penetrance to clinical disease in a large community cohort was partly explained by the cumulative effects of common genetic variants that influence iron measures in the general population. We showed that PRS for iron and transferrin saturation had the strongest associations with outcomes. We also showed that general population derived PRS for HH related conditions including liver diseases, diabetes and arthritis also modify penetrance to these respective diseases within p.C282Y homozygotes men and women. Therefore, including PRS in HH screening and diagnosis may help in estimating prognosis and treatment planning in p.C282Y homozygotes, especially those identified in population screening at younger ages before evidence of clinical endpoints could be present.

## Data Availability

Data are available to any bone fide researcher following application to UK Biobank (www.ukbiobank.ac.uk/register-apply). Access to UK Biobank was granted under Application Number 14631.

## Acknowledgements, Grant Support, and Disclosures

This work was generously funded by an award to DM by the Medical Research Council MR/S009892/1. DM and LP are supported by the University of Exeter Medical School. JA is supported by an NIHR Advanced Fellowship (NIHR301844).

The funders had no input in the study design; in the collection, analysis, and interpretation of data; in the writing of the report; or in the decision to submit the article for publication.

This research has been conducted using the UK Biobank Resource, under application 14631. The authors wish to thank the UK Biobank participants and coordinators for this unique dataset.

The authors would like to acknowledge the use of the University of Exeter High-Performance Computing (HPC) facility in carrying out this work.

All authors declare no conflicts of interest.

