## Supplementary Information (Figures) for "Penetrance of *HFE* haemochromatosis variants to clinical disease: polygenic risk score associations in UK Biobank"

### Supplementary Figure 1: Iron PRS associations with HH co-morbidities in UK Biobank males of European ancestry, stratified by *HFE* genotype

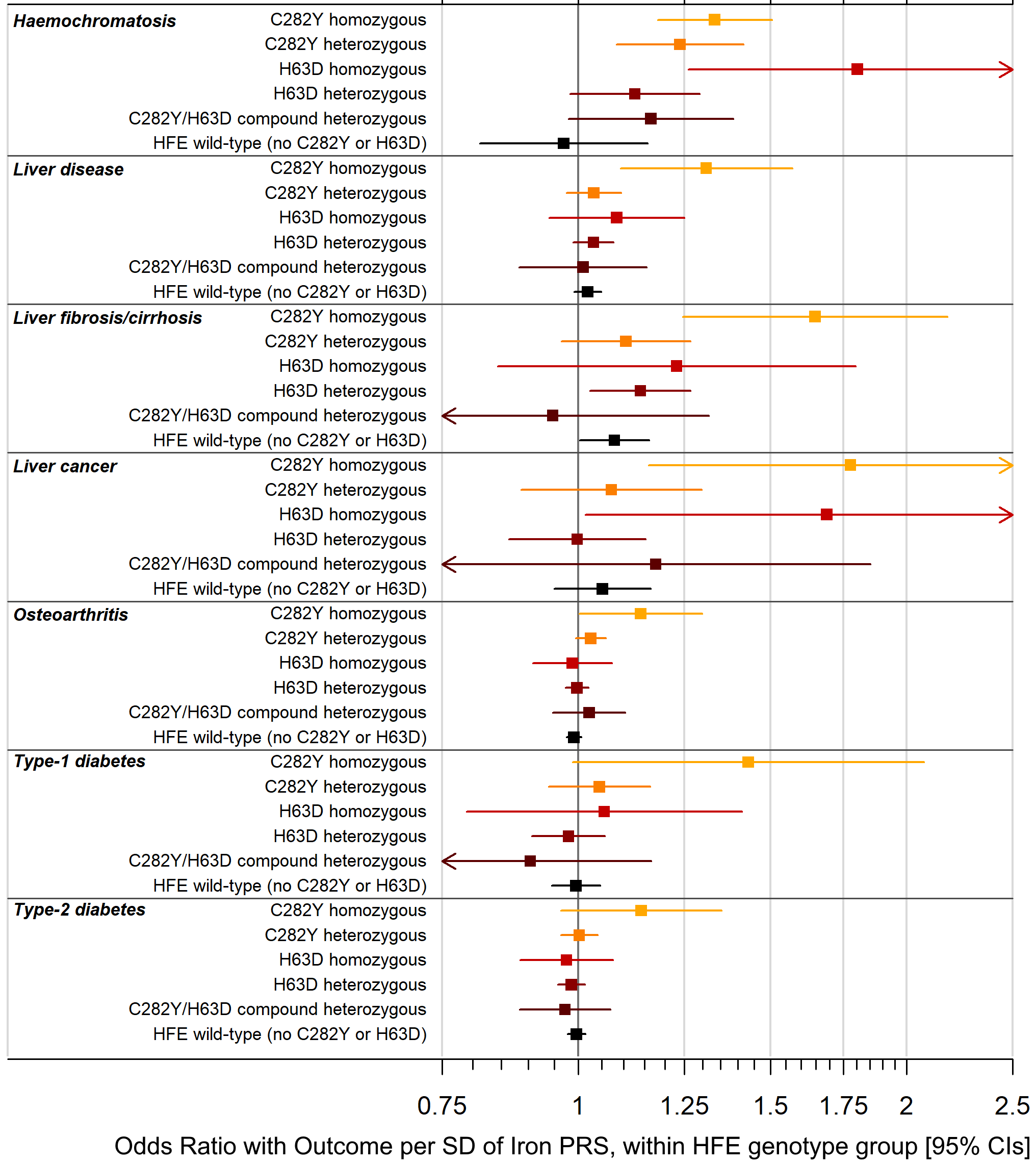
